# Repeated seroprevalence of anti-SARS-CoV-2 IgG antibodies in a population-based sample

**DOI:** 10.1101/2020.05.02.20088898

**Authors:** Silvia Stringhini, Ania Wisniak, Giovanni Piumatti, Andrew S. Azman, Stephen A. Lauer, Hélène Baysson, David De Ridder, Dusan Petrovic, Stephanie Schrempft, Kailing Marcus, Sabine Yerly, Isabelle Arm Vernez, Olivia Keiser, Samia Hurst, Klara M. Posfay-Barbe, Didier Trono, Didier Pittet, Laurent Gétaz, François Chappuis, Isabella Eckerle, Nicolas Vuilleumier, Benjamin Meyer, Antoine Flahault, Laurent Kaiser, Idris Guessous

## Abstract

**Background:** Assessing the burden of COVID-19 based on medically-attended case counts is suboptimal given its reliance on testing strategy, changing case definitions and the wide spectrum of disease presentation. Population-based serosurveys provide one avenue for estimating infection rates and monitoring the progression of the epidemic, overcoming many of these limitations.

**Methods:** Taking advantage of a pool of adult participants from population-representative surveys conducted in Geneva, Switzerland, we implemented a study consisting of 8 weekly serosurveys among these participants and their household members older than 5 years. We tested each participant for anti-SARS-CoV-2-IgG antibodies using a commercially available enzyme-linked immunosorbent assay (Euroimmun AG, Lübeck, Germany). We estimated seroprevalence using a Bayesian regression model taking into account test performance and adjusting for the age and sex of Geneva’s population.

**Results:** In the first three weeks, we enrolled 1335 participants coming from 633 households, with 16% <20 years of age and 53.6% female, a distribution similar to that of Geneva. In the first week, we estimated a seroprevalence of 3.1% (95% CI 0.2-5.99, n=343). This increased to 6.1% (95% CI 2.69.33, n=416) in the second, and to 9.7% (95% CI 6.1-13.11, n=576) in the third week. We found that 5-19 year-olds (6.0%, 95% CI 2.3-10.2%) had similar seroprevalence to 20-49 year olds (8.5%, 95%CI 4.99-11.7), while significantly lower seroprevalence was observed among those 50 and older (3.7%, 95% CI 0.99-6.0, p=0.0008).

**Interpretation:** Assuming that the presence of IgG antibodies is at least in the short-term associated with immunity, these results highlight that the epidemic is far from burning out simply due to herd immunity. Further, no differences in seroprevalence between children and middle age adults are observed. These results must be considered as Switzerland and the world look towards easing restrictions aimed at curbing transmission.

## Introduction

Although statistics on confirmed cases and deaths help in monitoring the dynamics of disease propagation, estimates remain largely unreliable when trying to understand the proportion of the population infected with SARS-CoV-2 for public health purposes^1^. For example, until recently most European countries, including Switzerland, did not have sufficient nasopharyngeal swabs available for RT-PCR screening of anyone suspected or at risk of infection with SARS-CoV-2. Generally, asymptomatic individuals, including those in at-risk categories, are not screened. As a result, the number of confirmed cases of SARS-CoV-2 infections is largely underestimated ^2^. In this context, seroprevalence surveys are of utmost importance for understanding the proportion of the population that has already developed antibodies against the virus and is potentially protected towards a new infection ^3^. As recommended by the World Health Organization (WHO), monitoring changes of seroprevalence over time is also crucial at the beginning of an epidemic to anticipate and plan an adequate public health response ^4^.

The canton of Geneva, Switzerland, reported its first confirmed COVID-19 case on February 26, with 5071 cases (10.15 per 1000 inhabitants) and 243 deaths as of April 30 ^5,6^. As in most countries, changing testing strategies over the course of the epidemics made it impossible to estimate the extent of the population that had been infected, while this information is crucial to plan evidence-based strategies to lift confinement measures. To assess the seroprevalence of anti-SARS-CoV-2 antibodies in this area, we initiated a survey in a representative sample of the population ^7^. To do so, we contacted subjects who already participated in the Bus Santé study (an annual health examination survey of a representative sample of the population of the canton) ^8^ and invited them along with their household members to participate to the SEROCoV-POP seroprevalence survey ^7^. Here we present results based on 1335 participants who took part to the study during the first 3 of 12 study weeks as they are likely to inform public health policy makers in Europe and beyond.

## Methods

### Study Design

Each week, ~1300 randomly selected previous participants of the Bus Santé study with an email address on file (n~10’072, **Figure S1**) were invited to participate in the SEROCoV-POP study by email, which provided a link to an online appointment booking system for a visit at one of two sites within the proceeding seven days. Eligible participants with a non-valid email address were contacted by phone to update their contact information. Since the third study week, each potential participant that hadn’t replied to the initial email invitation within 72h was reminded of the invitation by phone, in order to increase participation rate. Potential participants then received a confirmation by email, which included a link to a questionnaire and a consent form to complete at home and bring on the day of the study visit. During the visit, study staff discussed the study once again with participants to ensure informed consent. Participants were invited to bring all members of their household, aged 5 years and older, to join the SEROCoV-POP study. An electronic validation system allowed participants to declare that they weren’t in quarantine or isolation and didn’t present with symptoms compatible with COVID-19 at the moment of making the appointment, in which case they were encouraged to book at a further date. Participants considered vulnerable according to the Swiss Federal Office of Public Health criteria ^9^ (over 65 years old, diabetic, suffering from cardio-vascular or respiratory disease, immunocompromised, suffering from active cancer or with a BMI > 35 kg/m^2^) were asked to contact us directly by phone or email to book an appointment on time slots reserved explicitly for this population, in order to reduce risk of exposure to the virus. Inclusion criteria included people aged 5 years and older, whose primary residence was in the canton of Geneva. Over the first 3 weeks, 31% of invited eligible participants took part in the study, 16% refused to participate, and 53% have a pending status (waiting for booking appointment, being recalled, etc). We collected 6 mL of peripheral venous blood from each adult participant and 3 mL from each child less than 14 years old. The SEROCoV-POP study was approved by the Cantonal Research Ethics Commission of Geneva, Switzerland (CER16-363).

### Laboratory Analyses

We assessed anti-SARS-CoV-2-IgG antibodies using a commercially available enzyme-linked immunosorbent assay (Euroimmun AG, Lübeck, Germany # EI 2606-9601 G) targeting the S1-domain of the spike protein of SARS-CoV-2; sera diluted 1:101 were processed on a EuroLabWorkstation ELISA. An in-house validation study, using a set of sera from 176 pre-pandemic negative controls and 181 RT-PCR confirmed COVID-19 cases was conducted to estimate test performance and optimal thresholds for definitive results ^10^. Based on this validation study, which set thresholds to maximize specificity and inter-assay variability in test classification, we considered those with OD/CI >= 1.5 to be positive. This threshold resulted in a sensitivity of 86.2% and a specificity of 100%. For these analyses we treated all indeterminate results (>0.5 OD/CI <1.5) as negatives (Figure S1).

### Statistical Methods and Analyses

To estimate seroprevalence, we used a simple Bayesian regression model taking into account test performance, in addition to the age and sex of the population. In this framework, our goal is to estimate the true underlying seroprevalence, *p*, within the population:

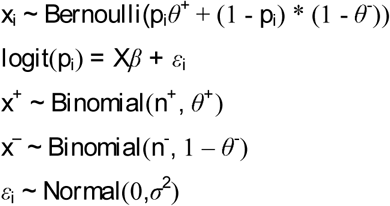

where *x_i_* is the result of the IgG ELISA for the *i*th person (*i* = 1, …, *N*). The sensitivity, *θ^+^*, is determined using *n*^+^ RT-PCR positive controls from the lab validation study, of which *x*^+^ tested positive. The specificity, *θ*^−^, is determined using *n*^−^ pre-pandemic negative controls, of which *x*^−^ tested positive. The probability of observing a diagnostic positive is a function of the true positive rate and the false negative rate with regards to the true underlying probability of seropositivity *p_i_*. This probability itself is a function of covariates *X*, which consists of sex, age categories, and week of study, and their coefficients *β* as well as an error term, *ϵ_i_* with variance *σ*^2^. We used naive priors on all parameters to allow for an exploration of the parameter space. We implemented this model in the Stan probabilistic programming language ^11^ and used the *rstan* package to run the model, analyse outputs. We ran 20,000 iterations with 4 chains and assessed convergence visually and using the R-hat statistic. While we did not account for household clustering specifically in this model, we developed a separate hierarchical model with a random intercept for household, including the modeling of test performance but with no covariates. We used this model to estimate the overall seroprevalence across the three weeks and compared to those from the model described above to understand the potential impact of not accounting for clustering within households.

To generate seroprevalence estimates adjusted to the age and sex distribution of the population, we sampled from the posterior for all age-sex strata proportional to the demographics in canton of Geneva. All estimates presented represent the mean of the posterior samples with the 2.5^th^ and 97.5^th^ percentiles of this distribution as the 95% confidence intervals (95%CI).

## Results

In the first three weeks of the study starting on April 6, we enrolled a total of 1335 individuals in the SEROCoV-POP survey (Figure S2). Included participants came from 633 different households, with 53.6% being female, and 16% being under 20 years of age (**Table 1**), displaying a similar distribution to that of the overall Geneva population ^12^. While 246 individuals participated alone to the study, 193 participants came with one other household member, 97 participants with two household members, and 97 participants with three or more household members.

**Table 1:** Overview of sample size and seroprevalence estimates by week, sex and age. Note that p-values are Bayesian p-values following Gelman et al. ^18^.

|  | **n** | **positive** | **seroprevalance** | **p-value** |
| --- | --- | --- | --- | --- |
| *Age* |  |  |  |  |
| 5-19 | 214 | 13 (6.1%) | 6.0, 95% CI (2.3-10.2) | 0.12 |
| 20-49 | 538 | 45 (8.4%) | 8.5, 95% CI (4.9-11.7) | - |
| 50+ | 583 | 25 (4.3%) | 3.7, 95% CI (0.9-6.0) | <0.001 |
| *Sex* |  |  |  |  |
| Female | 715 | 40 (5.6%) | 5.6, 95% CI (3.1-8.1) | - |
| Male | 620 | 43 (6.9%) | 6.9, 95% CI (3.3-9.9) | 0.24 |
| *Week* |  |  |  |  |
| Week 1 | 343 | 11 (3.2%) | 3.1, 95% CI (0.2-5.9) | 0.03 |
| Week 2 | 416 | 22 (5.3%) | 6.1, 95% CI (2.6-9.33) | - |
| Week 3 | 576 | 50 (8.7%) | 9.7, 95% CI (6.1-13.1) | 0.03 |

In the first week, we estimated an overall seroprevalence of 3.1% (95% CI 0.2-5.9). This figure increased to 6.1% (95%CI 2.6-9.33, n=416) in the second week, and to 9.7% (95% CI 6.1-13.1, n=576) in the third week, mirroring the increase in confirmed cases in the weeks before the survey (**Figure 1**). While there were no meaningful differences in seroprevalence between men and women (**Table 1**), we found that 5-19 year olds (6.0%, 95% CI 2.3-10.2%) had a similar seroprevalence to 20-49 year olds (8.5%, 95%CI 4.9-11.7), whereas seroprevalence among those 50 and older (3.7%, 95% CI 0.9-6.0, p<0.01) was significantly lower.

**Figure 1.**
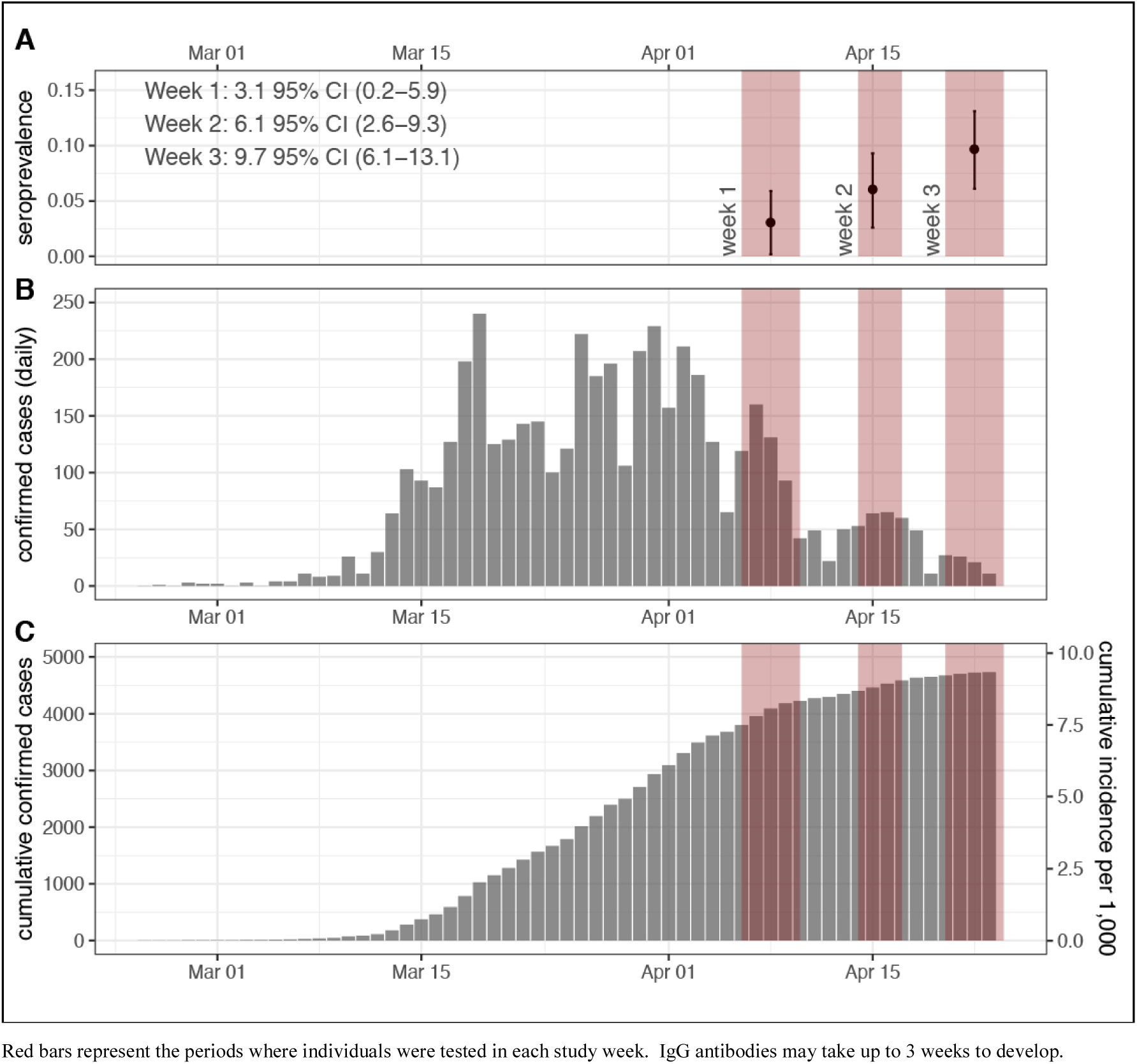
Seroprevalence estimates and 95% confidence interval (CI)s for each week of the survey (A), daily confirmed cases reported in Geneva (B) and cumulative case counts per day and cumulative incidence rate of confirmed COVID-19 (C).

As a sensitivity analysis, we estimated seroprevalence among the subset of participants who were originally enrolled in our previous representative surveys (**Table 2**). Within this subset, we observed similar seroprevalence as in the overall study population, with higher estimates in younger individuals (20-49 years vs. 50+ years), and with increasing seroprevalence estimates from the first to the third week of the study.

## Discussion

The preliminary results of this study provide an important benchmark to assess the state of the epidemic. With an estimated 48’500 people having developed antibodies (9.7% of 500’000 inhabitants) while 4741 cases were confirmed on April 24^th^ ^5^, we observe that there are roughly 10 infections for every COVID-19 confirmed case in Geneva, reflecting the variability in disease severity, testing practices and care-seeking behaviors. Further, we show that three weeks after the peak of confirmed cases, only 1 in 10 people has developed antibodies against SARS-CoV-2, even in one of the more heavily affected areas in Europe ^13^. Thus, assuming that the presence of IgG antibodies measured in this study is at least in the short-term associated with immunity, these results highlight that the epidemic is far from burning out simply due to herd immunity. Given the time to development of IgG antibodies (1-3 weeks), we expect to continue seeing significant increases in seroprevalence over the coming weeks ^14^.

We found no differences in seroprevalence between children/young-adults (5-19 years old) and middle age adults, with those older than 50 years having a significantly lower seroprevalence than 20-49 year olds. Although children present typical COVID-19 symptoms far less frequently than adults, these results provide support to emerging research showing that they indeed get infected at similar rates ^15^. This should be considered in view of increased concerns about severe inflammatory syndromes in children that could be COVID-19-related ^16^, and of the worldwide debate around opportunity and modality of school re-openings. While the sample size of older adults was small, the lower seroprevalence estimates in this group suggest that targeted efforts to reduce social mixing of elderly people with others may have succeeded. However, it remains possible that, because of an age-related compromised ability to generate adaptive immune responses, the elderly develop a lower IgG response after infection, something that needs further investigation ^17^.

To our knowledge, this is the first study to perform a large-scale seroprevalence survey of anti-SARS-CoV-2 IgG antibodies where participants were selected from a representative sample of the general population. It is also unique in its repeated nature allowing the study of immunity dynamics in both adult and children populations. Our population-based design as well as the fact that we informed participants that individual results were not going to be disclosed until the end of the study, mitigate selection bias. This study applies advanced statistical methods accounting for demographic structure and imperfect diagnostic test to estimate seroprevalence in the overall population while capturing uncertainty in the estimates.

Our study also has some important limitations to acknowledge. First, the primary analyses include randomly selected participants as well as of members of their households, making this sample not entirely randomly selected. We attempt to adjust for some aspects of this through poststratification within our statistical model. Further, in sensitivity analyses we estimated the seroprevalence for just the original and found that it is similar to that of the full sample. Second, we were able to account for clustering within households only for overall prevalence pooling the three weeks, as there was not enough data to account for age, sex, and household clustering all together within this framework. The use of a random effects model to account for household clustering did not change pooled results. Finally, the recruitment of participants by email might exclude non tech-proficient individuals or people without access to technology; another survey specifically targeting vulnerable populations (socially and clinically) is ongoing.

Over the next weeks, we will continue monitoring weekly seroprevalence in the general population and will be able to provide more refined analysis on symptomatology and other socio-demographic data in relation to immunological status. Yet, a preliminary presentation of these results is deemed to be necessary to timely inform global policy makers on how to adapt planning of the next phases. Public sharing of our protocol can also help the global academic community to implement serosurveys in their areas. For at least one year, we will follow up participants via an online digital platform and repeated serological/RT-PCR testing in order to assess COVID19 incidence in the population. These longitudinal results will also inform us on the dynamics of immunity which remain debated at this stage.

Our results highlight that as hospitalizations have reduced in Geneva and other similar locations throughout the world, the immunologic landscape has not changed greatly since the onset of the pandemic, with the vast majority of people having no evidence of past infection. As the world develops plans to find a new balance between minimizing the direct impacts of COVID-19 on those infected and the indirect effects on all of society, serologic studies such as this are critical for providing new insights about transmission and the otherwise hidden immunologic state of the population.

## Data Availability

Data can be made available upon reasonable request at the completion of the study through contacting the study PI.

## Acknowledgments

This study would not have been possible without the instrumental and passionate contribution of the staff of the Unit of Population Epidemiology of the HUG Primary Care Division (Natacha Noel, Caroline Pugin, Jane Portier, Barinjaka Rakotomiaramanana, Leila Botudi, Natalie Francioli, Paola d’Ippolito, Chantal Martinez, Francesco Pennacchio, Benjamin Emery, Zoé Waldman, Magdalena Schellongova, Prune Collombet), of the team of the Division of Laboratory Medicine (Géraldine Poulain and Pierre Lescuyer), of the HUG staff affected to our unit during the COVID emergency (Lilas Salzmann-Bellard, Mélanie Seixas Miranda, Yasmina Malim, Acem Gonul, Odile Desvachez) and finally without the invaluable work of the medical students who have invested their time and energy in this project (Jonathan Barbolini, Eugénie de Weck, Natacha Michel, Emmanuelle Mohbat, Irine Sakvarelidze, Céline Eelbode, Sultan Bahta, Céline Dubas, Lina Hassar, Melis Kir, Hugo-Ken Oulevey, Kourosh Massiha, Manon Will, Natacha Vincent, Fanny Lombard, Alioucha Davidovic, Benoit Favre, Amélie Mach, Eva Marchetti, Sophie Cattani, Joséphine Duc, Julie Guérin, Soraya Maret, Francesca Hovagemyan, Antoine Daeniker, Rebecca Buetzberger). We also thank Professor Nicola Low for her insight and help in designing the study, as well as all the participants of the Bus Santé Study and their household members.

## Funding

This study was funded by the Swiss Federal Office of Public Health, The Swiss School of Public Health, the Fondation de Bienfaisance du Groupe Pictet, the Fondation Ancrage, the Fondation Privée des HUG, the Center for Emerging Viral Diseases. The funders had no role in study design, data collection and analysis, decision to publish, or preparation of the manuscript.

## Author’s contribution

SS, AW, AF, LK and IG conceived the study. SS and IG drafted the first version of the manuscript. AA, AW, DP contributed to drafting sections of the manuscript. AA, SL and GP performed data analyses. SY, IAV, IE, NV, BM, LK performed lab analysis. GP, HB, AA, SL, DDR, DP, StS, KM, OK, SH, KPB, DT, DP, LG, FC participated in the study design and helped to draft the manuscript. All authors contributed to the interpretation of data, and read and approved the final manuscript.

## Conflicts of interest to disclose

No author had conflicts of interest to disclose.

**Figure S1.**
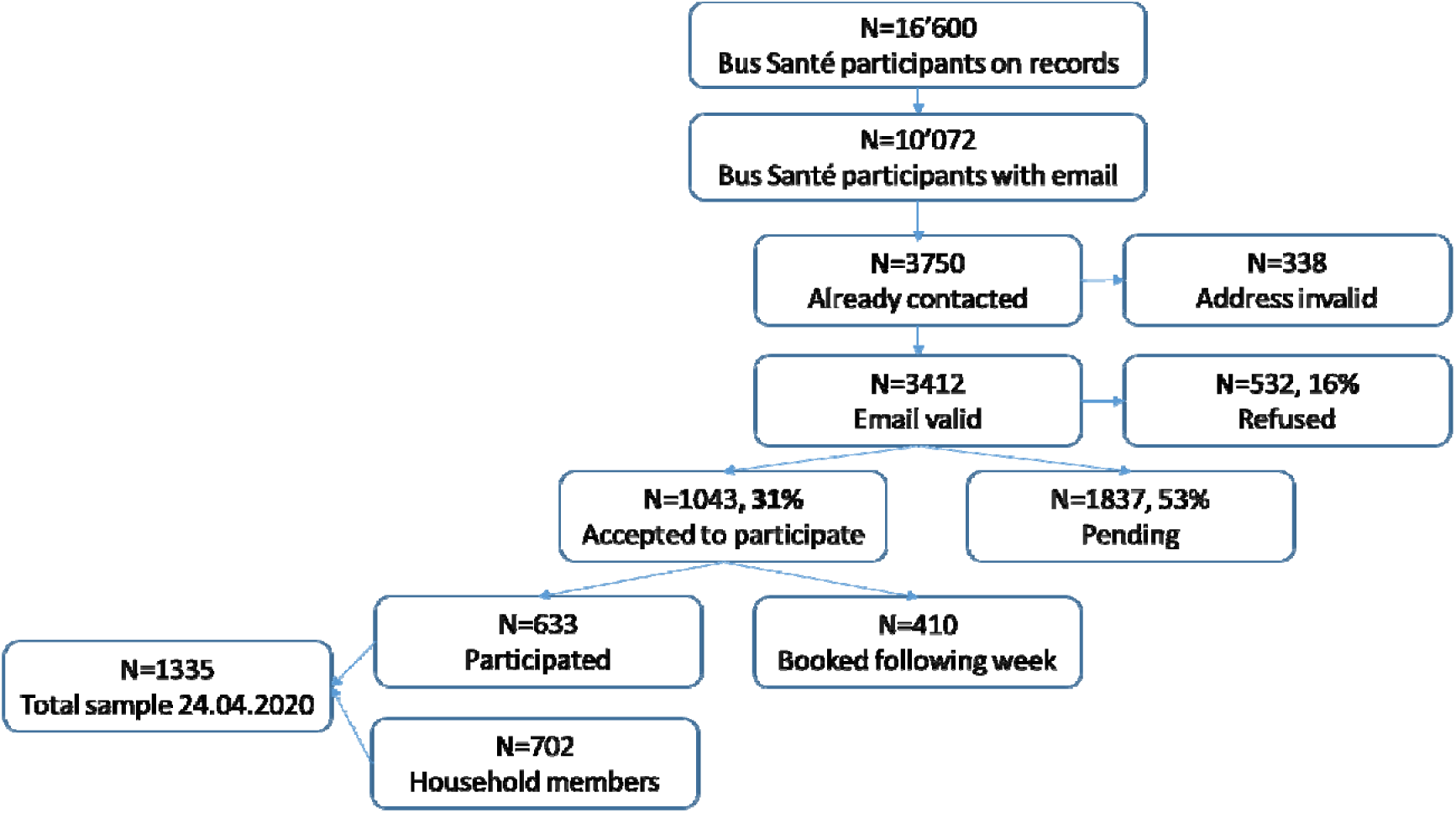
Flow chart of inclusion in the study.

**Table S1:** Overview of sample size and seroprevalence estimates by week, sex and age (Bus Santé randomly selected participants only – other household members were excluded).

|  | n | positive | seroprevalance | p-value |
| --- | --- | --- | --- | --- |
| *Age* |  |  |  |  |
| 20-49 | 253 | 2222 (8.7%) | 9.5, 95% CI (5.3-14.0) | - |
| 50+ | 375 | 15 (4.0%) | 3.7, 95% CI (1.0-6.4) | 0.00 |
| *Sex* |  |  |  |  |
| Female | 340 | 1616 (4.7%) | 5.7, 95% CI (2.8-9.1) | - |
| Male | 288 | 21 (7.3%) | 8.0, 95% CI (3.7-12.5) | 0.19 |
| *Week* |  |  |  |  |
| Week 1 | 156 | 4 (2.6%) | 3.0, 95% CI (0.1-7.2) | 0.02 |
| Week 2 | 190 | 12 (6.3%) | 8.3, 95% CI (4.1-13.4) | - |
| Week 3 | 282 | 21 (7.4%) | 9.0, 95% CI (4.5-13.7) | 0.39 |

**Figure S2.**
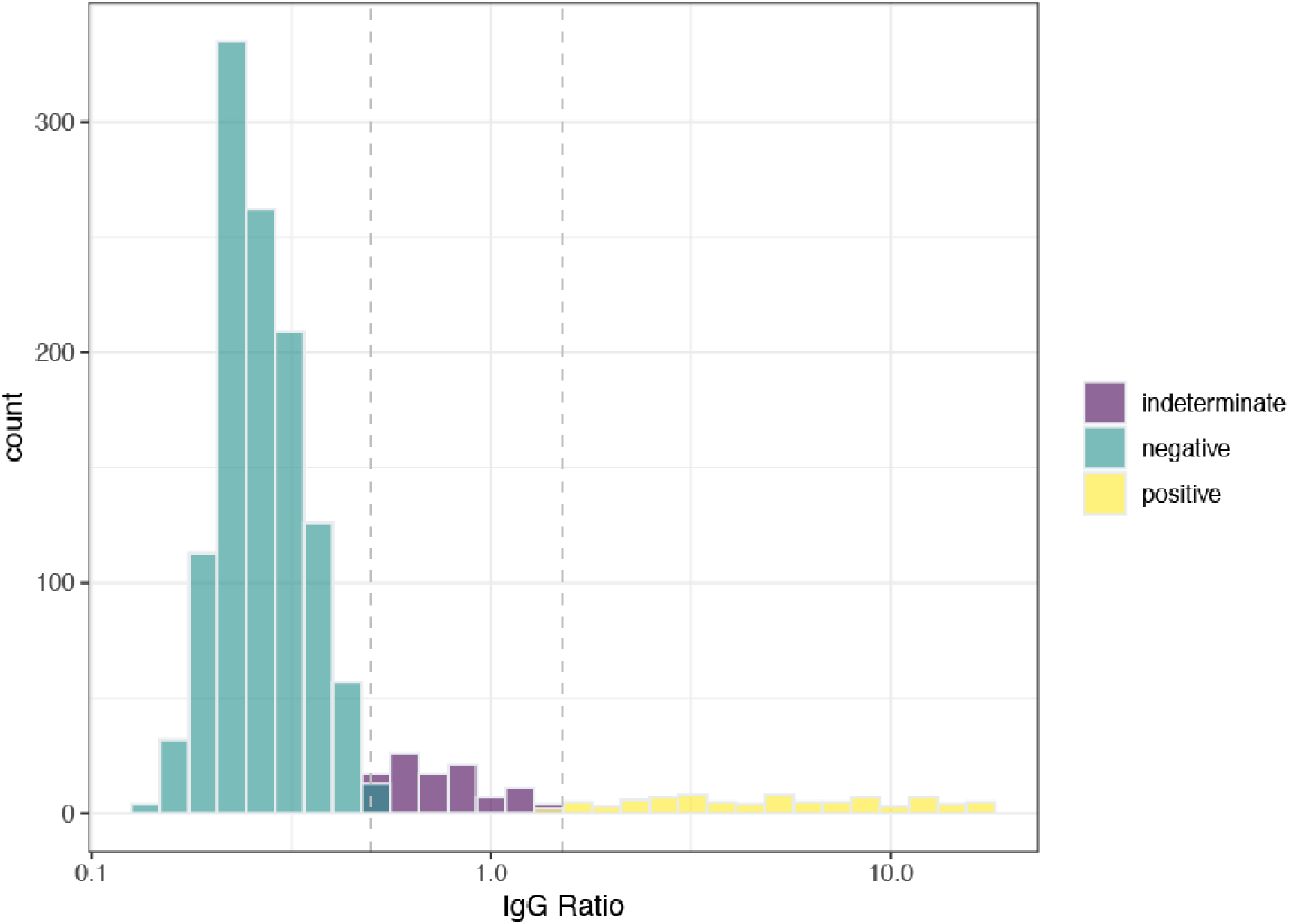
Density of ELISA ratios for all participants colored by result interpretation. All indeterminates were considered negative for these analyses.

## Notes

### Competing Interest Statement

The authors have declared no competing interest.

### Clinical Protocols

https://www.covicare24.com/s/Protocole_SEROCOV-POP3003.pdf

